# Sputum microbiota profiles of treatment-naïve TB patients in Uganda before and during first-line therapy

**DOI:** 10.1101/2020.04.24.20078246

**Authors:** David Patrick Kateete, Monica M Mbabazi, Faith Nakazzi, Fred A Katabazi, Edgar Kigozi, Willy Ssengooba, Lydia Nakiyingi, Sharon Namiiro, Alphonse Okwera, Moses L Joloba, Adrian Muwonge

## Abstract

There is limited information on microbiota dynamics in tuberculosis (TB) in Africa. Here, we investigated changes in microbiota composition, abundance, co-occurrence and community remodelling relative to clinical parameters, among treatment-naïve pulmonary TB patients at Mulago National Referral Hospital in Kampala, Uganda. We sequenced 205 sputum samples from 120 patients before initiating anti-TB therapy (baseline) and during treatment follow-up (at months 2 and 5). A total of 8.6 million high quality sequences were generated, yielding 8,180 operational taxonomic units (OTUs), 18 phyla and 333 genera. A sputum sample on average generated 44,992 sequences, yielding 6,580 OTUs, 4 phyla and 36 genera. The sputum microbiota core comprised of 34 genera and it was remarkably stable during treatment. Month 2 was characterized by a significant mean reduction in core microbiota biomass, limited variance changes and general lack of entropy. However, variance and entropy recovered at month 5. Co-occurrence patterns were predominated by accessory genera at baseline but their abundance significantly reduced during treatment. Our findings reveal discernible sputum microbiota signals associated with first-line anti-TB therapy, with potential to inform treatment response monitoring in developing countries.

## Introduction

Tuberculosis (TB) is a persistent public health problem and one of the top 10 causes of death worldwide^1^. Nearly half a million new TB cases have been reported in Uganda since 2010^2^, and the TB incidence in the country surpassed that of HIV-infection in 2016^3^. While the introduction of the Xpert MTB/RIF assay revolutionized the diagnosis of TB globally^4^, treatment still hinges on long treatment regimens i.e. 6 to 24 months depending on whether treating drug susceptible TB or drug resistant TB^5^. The standard first-line treatment regimen comprises of an intensive phase of 2 months treatment with isoniazid, rifampicin, pyrazinamide and ethambutol, followed by a continuation phase of 4 months treatment with isoniazid and rifampicin^6,7^. After initiating therapy, sputum microscopy for identification of mycobacteria (in form of acid-fast bacilli, AFB) or sputum culturing for *Mycobacterium tuberculosis* growth, are regularly done during the treatment period usually at months 2 and 5 to monitor treatment response. Attaining sputum sterilisation, also known as sputum smear-conversion or sputum culture-conversion (i.e. from AFB/culture positive to AFB/culture negative) at months 2 or 5 after initiating therapy is a known cardinal index of treatment success. In fact, the World Health Organization (WHO) guidelines state that “a patient whose sputum was AFB smear-positive or culture-positive at the beginning of therapy but becomes smear-negative or culture-negative in the last month of treatment and on at least one previous occasion is declared cured” ^6,8^. Despite the importance of sputum-smear / sputum-culture conversion in monitoring treatment response^9^, they have low sensitivity and in low-income countries culture is not routinely done. However, as sputum is the cornerstone in diagnosing TB, holistic identification of microbiological factors in sputum and their association with treatment response could unravel new ways in which TB diagnosis or treatment response monitoring can be improved^7^.

Microbiota are ‘microorganisms’ –bacteria, fungi, protozoa and viruses that live on the skin and mucosa of humans and other mammals. Their role in induction, maintaining, disrupting and modulation of the immune response has recently come into focus with the advent of the human microbiome project^10^. Microbiota also exist in the lung^11^, the predilection site for the TB causative agent *M. tuberculosis*, and perhaps influence its behaviour in a variety of ways e.g. signalling^11-13^. Therefore, sound understanding of the microbiota in TB is necessary given their emerging importance in human and animal health^11,14^. In this context, microbiota profiling in pulmonary TB can advance our knowledge of TB pathogenesis and unravel new ways in which TB diagnostics might be improved^7,11-13^. While microbiota/microbiome studies in TB have advanced^11,12,15,16^, there is a general lack of knowledge in sub-Saharan Africa^11^ where the TB burden is greatest. The aim of this study was to investigate the microbiota profiles among treatment-naïve TB in Kampala, Uganda, and the impact of first-line anti-TB therapy on the microbiota, using sputum as proxy for the lung environment^17^. We describe sputum microbiota changes during critical transitions of anti-TB therapy: Pre-treatment (baseline) and treatment response follow-up at months 2 and 5.

## Results and Discussion

### Demographics

This study enrolled 120 treatment-naïve pulmonary TB patients at Mulago National Referral Hospital in Kampala Uganda, in the period between 2016 and 2018. Two hundred and five sputum samples were collected from the patients and sequenced –120 at baseline, 44 at month 2, and 41 at month 5 (**Figure 1**). **Table 1** summarises the clinical and demographic characteristics of the patients. The mean age was 33 years; majority were male, residents of Kampala and Wakiso districts (**Supplementary Figure S1** online).

**Table 1:** Clinical and demographic characteristics of pulmonary TB patients enrolled (n=120)

| Variable | Patients (%) | Proportion by gender (%) |  |
| --- | --- | --- | --- |
|  |  | Female | Male |
| **HIV status** |  |  |  |
| Positive | 26 (21.7) | 12 (46.2) | 14 (53.8) |
| Negative | 94 (78.3) | 32 (34) | 62 (66) |
| **Chest X-ray** |  |  |  |
| Cavities | 66 | 25 | 41 |
| No cavities | 37 | 10 | 27 |
| **Medication** |  |  |  |
| HIV-positive on ART | 16 (60) | 8 (0.5) | 8 (0.5) |
| History of antibiotic use | 22 (18.3) | 8 (36.4) | 14 (63.6) |
| Others* | 23 (19.2) | 11 (47.8) | 12 (52.2) |
| **Comorbidity** |  |  |  |
| High blood pressure / hypertension | 10 (8.3) | 2 (20) | 8 (80) |
| Cigarette smokers | 26 (21.7) | 1 (3.8) | 25 (96.2) |
| Alcohol consumers | 40 (33.5) | 11 (27.5) | 29 (72.5) |
| **BMI** |  |  |  |
| Low ($\leq 18$) | 64 (53.3) | 21 (32.8) | 43 (67.2) |
| Medium ($\geq 19$-25) | 52 (43.4) | 19 (36.5) | 33 (63.5) |
| High ($\geq 26$-29) | 4 (3.3) | 4 (100) | 0 |
| **Education** |  |  |  |
| Illiterate | 7 (5.8) | 4 (57.1) | 3 (42.9) |
| Primary level | 27 (22.5) | 12 (44.4) | 15 (55.6) |
| Secondary level (O-levels) | 86 (71.7) | 29 (33.7) | 57 (66.3) |
| **District of residence** |  |  |  |
| Kampala | 63 (52.5) | 26 (41.3) | 37 (58.7) |
| Wakiso | 40 (33.3) | 14 (35) | 26 (65) |
| Others** | 17 (14.2) | 4 (23.5) | 13 (76.5) |
| **Ancestry** |  |  |  |
| Bantu | 100 (83.3) | 37 (37) | 63 (67) |
| Non-Bantu | 20 (16.7) | 7 (35) | 13 (65) |
| **Employment status** |  |  |  |
| Employed | 97 (80.8) | 34 (35.1) | 63 (64.9) |
| Not employed | 23 (19.2) | 10 (43.5) | 13 (56.5) |
| **CFM** |  |  |  |
| No AFB | 19 (15.8) | 9 (47.4) | 10 (52.6) |
| Scanty | 20 (16.7) | 6 (30) | 14 (70) |
| 1+ AFB | 12 (10) | 4 (33.3) | 8 (66.7) |
| 2+ AFB | 30 (25) | 12 (40) | 18 (60) |
| 3+ AFB | 39 (32.5) | 13 (33.3) | 26 (66.7) |
| **Sampling point** |  |  |  |
| Baseline (before treatment) | 120 (100) | 53 (44.2) | 67 (55.8) |
| 1<sup>st</sup> treatment follow up (2 months after) | 44 (36.7) | 15 (34.1) | 29 (65.9) |
| 2<sup>nd</sup> treatment follow up (5 months after) | 41 (34.2) | 16 (39) | 25 (61) |

- \*Refers to medicines other than antibiotics & antiretroviral (ARVs) - \*\*Refers to participants staying in districts other than Kampala & Wakiso. Difference in sums was due to sequences removed during filtering. - ART, antiretroviral therapy; AFB, acid fast bacilli; BMI, body mass index; CFM, conventional fluorescence microscopy; COPD, chronic obstructive pulmonary disease

**Figure 1:**
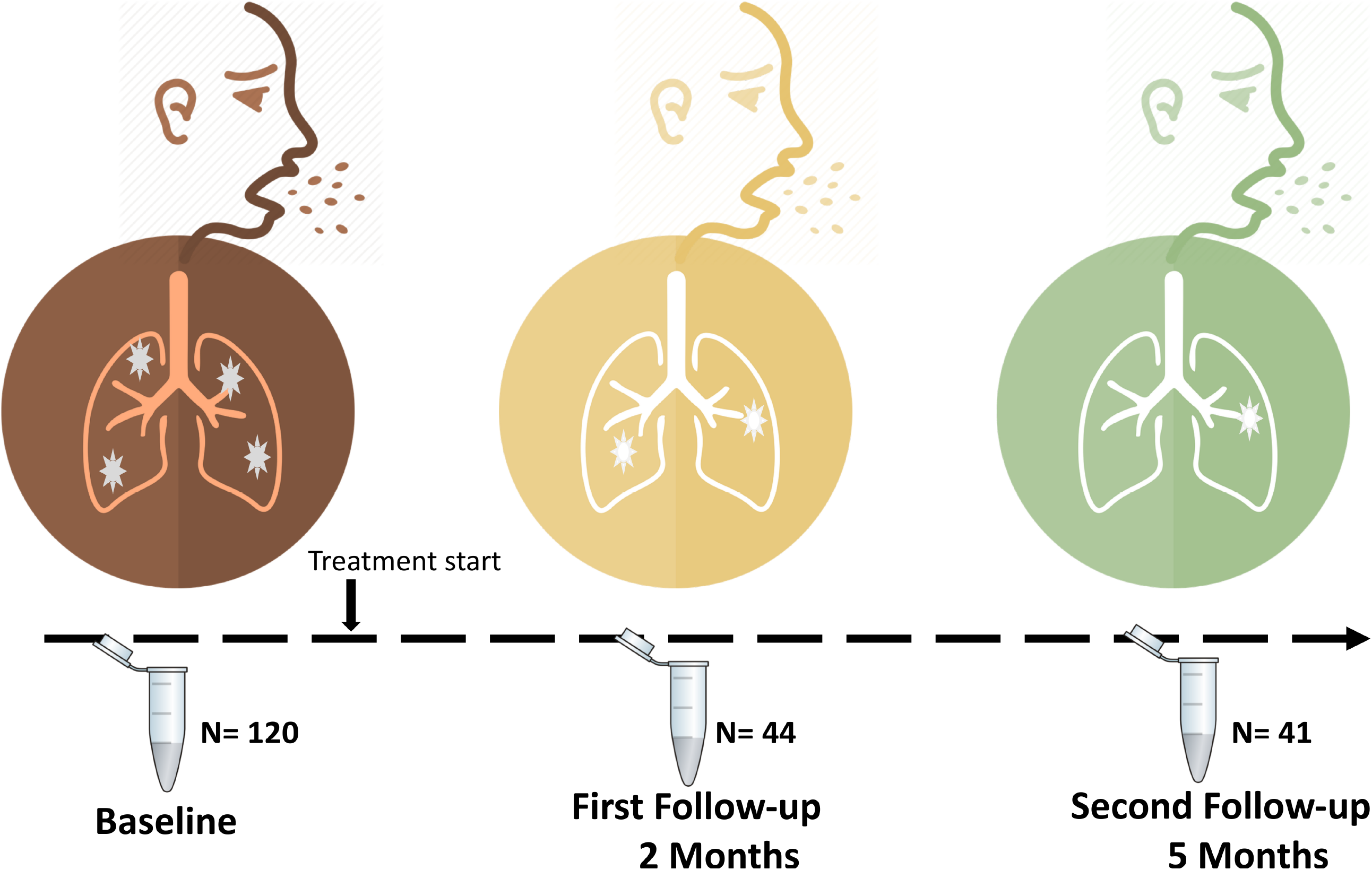
Study design and layout in this study. Dark red, orange and green icons depict Baseline (before initiating therapy), Months 2 and 5 posttreatment commencement, respectively. A total of 120 pulmonary TB patients were enrolled at baseline but on follow-up, we screened 44 and 41, respectively.

### Sputum microbiota structure of treatment-naïve TB patients

High-throughput sequencing of the variable region of the *16S rRNA* gene generated a total of 9,316,821 sequence reads from the 205 sputum samples. After filtering and quality control, we retained 8,638,640 sequences representing 192 samples i.e. 13 samples were removed due to low sequence quality. The retained high quality sequences yielded 8,180 OTUs, 18 phyla and 333 genera. A sputum sample on average generated 44,992 sequences, yielding 6,580 OTUs, 4 phyla and 36 genera. Six hundred and seventeen OTUs and 91 genera were shared between patients across the sampling points i.e. baseline, months 2 and 5 (**Supplementary Figure S2** online). Bacteroidetes, Firmicutes, Proteobacteria, Fusobacteria and Actinobacteria were the most predominant phyla accounting for nearly 95% of the sputum microbiota composition. We observed that negative sputum samples on Löwenstein-Jensen (LJ) culturing produced more sequence reads (**Supplementary files Table S1** and **Figure S3** online) however, the *Mycobacterium* sequences detected matched the expected outcomes with routine TB diagnostic methods i.e. positive likelihood ratio of 0.71 (95% CI=0.59, 0.85). While sputum microbiota of pulmonary TB patients has been investigated before and researchers provided key insight into its composition and diversity, majority of the studies were conducted in Asia^11,16,18,19^. Our study generated nearly 16 times the number of OTUs reported by investigators in Asia^15,16^, probably because of the latest amplicon sequencing technology we used (i.e. MiSeq/illumina Inc. vs. 456/Pyrosequencing in previous studies). Conversely, these differences may signify existence of high sputum microbiota diversity among TB patients in Uganda: Indeed, the microbiota diversity observed in this study accounts for nearly 48% of the diversity in the human oral microbiome database http://www.homd.org.

#### Alpha and beta diversity

On average, a sputum sample from a treatment-naïve TB patient had a Shannon diversity index of 3.8 and this was not affected by HIV status, contradicting some reports from Asia^16,20^. Generally we observed no difference with respect to sampling points (i.e. baseline, months 2 & 5) at alpha diversity level on indices like richness, Simpson and Shannon (**Figure 2, panel A**). However for beta diversity, we observed distinct clustering of patients when we analysed treatment follow-up samples relative to baseline samples (before initiating therapy) **Fig**.**2 (panel B)** suggesting occurrence of sputum microbial signals during treatment that could be exploited for insight into improving treatment response monitoring. Clinical and lifestyle variables in this study explained approx. 14% of the microbial variance. Most of this variance was attributed to treatment follow up samples (months 2 & 5), which again highlights the potential utility of sputum microbiota in monitoring treatment response^21^. It is noteworthy that the total explained variance observed was similar to that estimated in PERMANOVA (permutational multivariate analysis of variance, see Methods). The relationship between effect-sizes of clinical variables and Bray-Curtis changes, weighted and unweighted uniFrac distances is shown in **Supplementary files Fig**.**S4** and **Table S3** online (i.e. colony forming units, 1.6%, 1.6%, 1.1%; body mass index, 1.1%, 1.0%, 0.8%; sampling point, 1.4%, 2.3%, 2.0%; nutritional status, 0.6%, 0.5%, 0.7%; and smoking, 0.5%, 0.7%, 2.7%). In this PERMANOVA analysis these variables cumulatively explained 11-14% of the variance across beta diversity indices. Since the largest proportion of explained variance was attributed to sampling point and sputum smear staining (**Fig.2 panel B**), it is likely that additional sputum microbial factors that change with treatment do occur.

**Figure 2:**
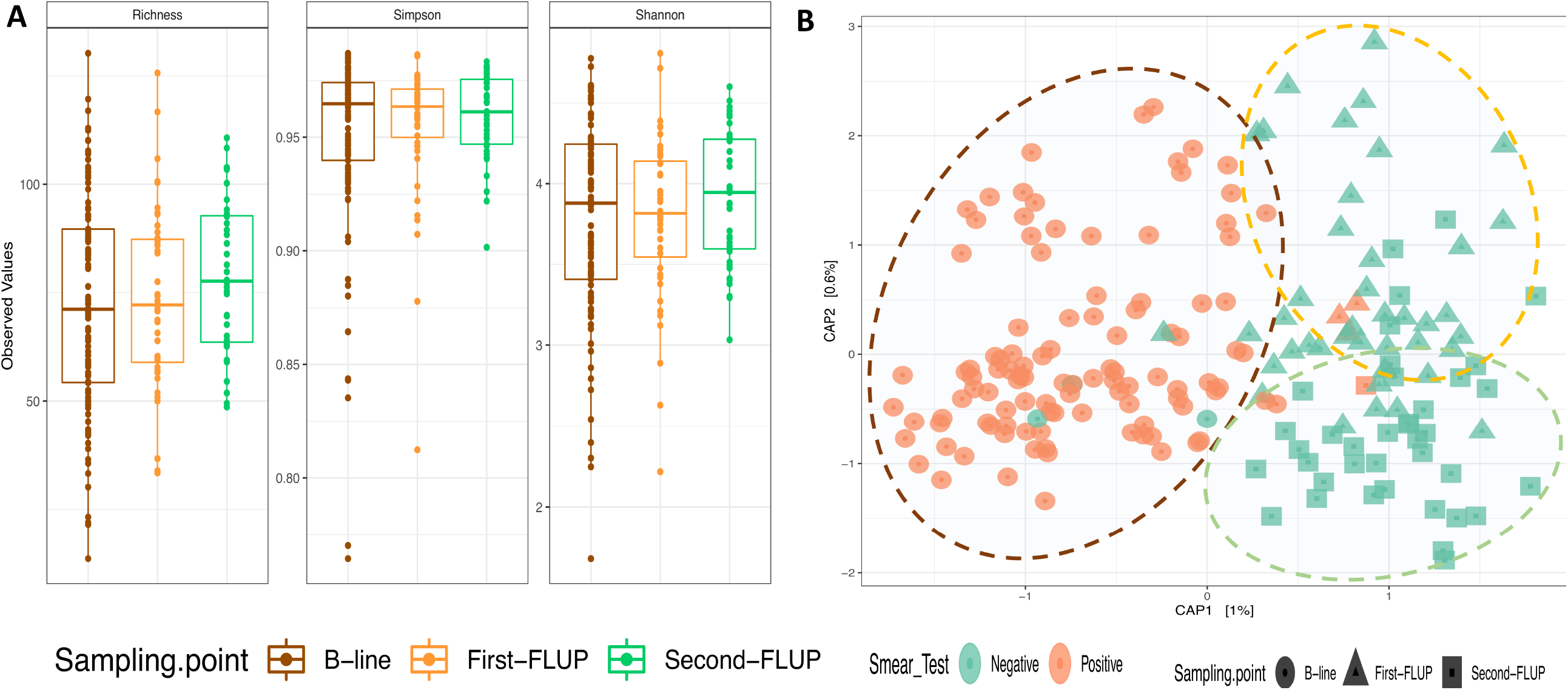
Microbial diversity analysis during first-line anti-TB treatment. Panel A shows Alpha diversity analysis for indices like Richness, Simpson and Shannon. Panel B shows Beta diversity analysis based on constrained ordination of the Bray-Curtis distances. The green and colors represent positive and negative outcomes on routine smear diagnosis. B-line, Baseline before initiating therapy; Fist-FLUP, First follow up (month 2); Second FLUP, Second follow up (month 5).

### Taxonomy characteristics during treatment: Accessory microbiota fluctuates while core microbiota is stable

To unravel the effect of anti-TB treatment on the microbiota, we characterised and tracked the size and composition of the ‘core’ and ‘accessory’ microbiota in sputum samples collected during treatment follow-up relative to baseline samples (**Figures 3-5**). By ‘core microbiota’ we refer to genera present in eighty percent of samples at a given sampling point^22^, in this case baseline, months 2 and 5; ‘accessory microbiota’ refers to the difference between the core and richness and it provides insight into transient microbiota. Changes in abundance of genera characterised as core microbiota at each sampling point are shown in **Figures 3** and **4**; interestingly, the core size was remarkably stable with an average of 34 genera over the sampling period. It is noteworthy that *Mycobacterium*’s membership of the core was restricted to baseline samples before initiating therapy (**Figure 3**). The core comprised of genera like *Streptococcus, Veillonella, Neisseria, Fusobacterium, Lachnoanaerobaculum, Atopobium, Peptostreptococcus* and *Leptotrichia*. Of these, *Streptococcus* was the most abundant but unlike *Veillonella*, its abundance was consistent before and after initiating therapy. Conversely, genera like *Neisseria* showed a steady increase over time. To track changes in the core, we identified 31 genera whose core membership was consistent across the three sampling points (**Figure 5**). The 31 genera are the same as genera characterised as normal flora; this core and normal flora as a proportion of sputum microbial richness was also stable across sampling points (**Figure 5**). In contrast, by month 2 after initiating treatment, the proportion of *Mycobacterium* dramatically fell while that of accessory microbiota fluctuated (**Figure 5**). It is worth noting that the proportion of accessory microbiota was at its highest at baseline and reduced at month 2 after initiating treatment.

**Figure 3:**
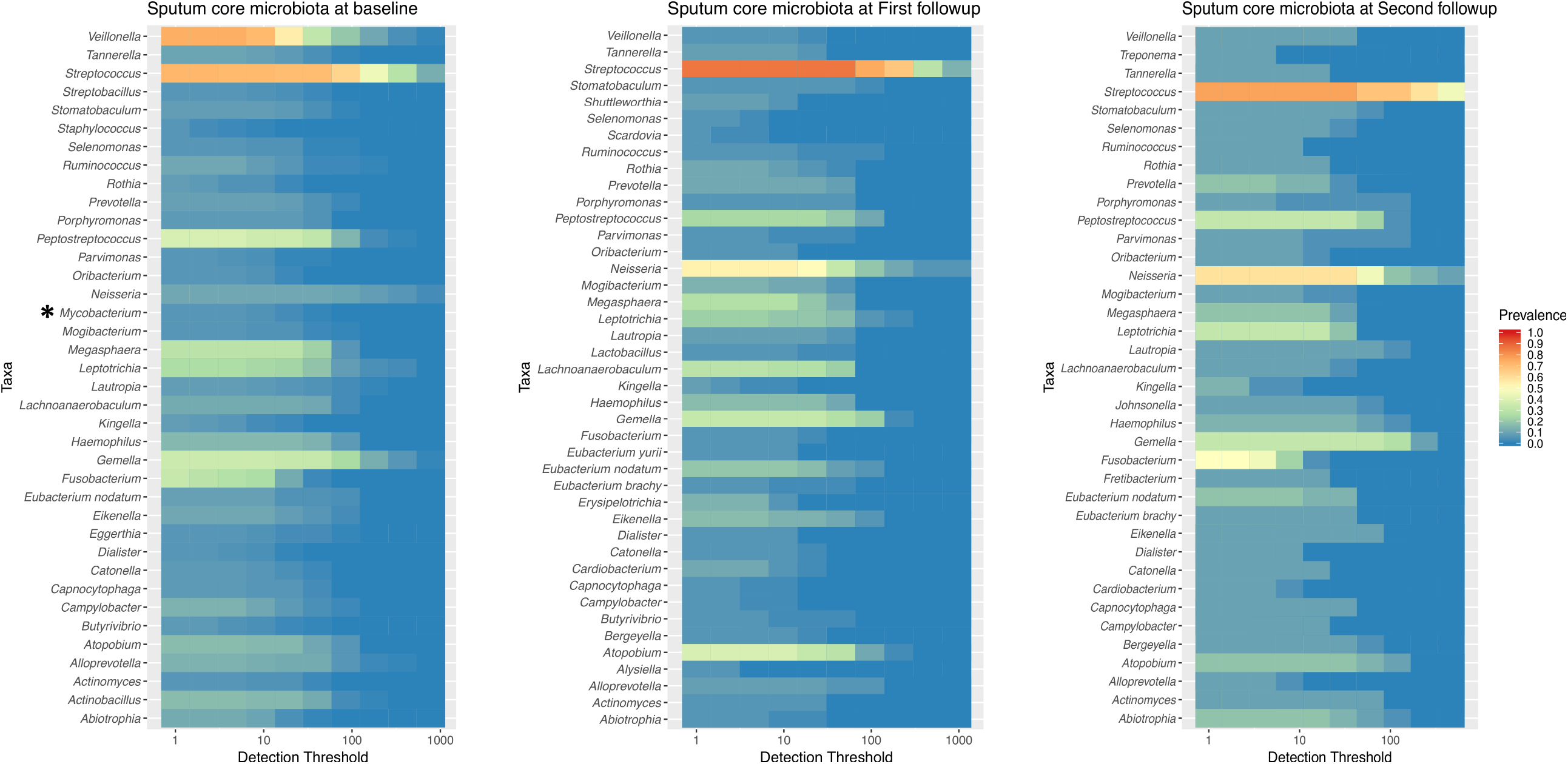
Taxonomic characteristics of the microbiota during anti-TB therapy. **Panel 3** depicts composition of the core genera during treatment as defined by the QIIME-2 microbiome package. Note that the genus *Mycobacterium* is a member of the core before initiating therapy but not during/after treatment.

**Figure 5:**
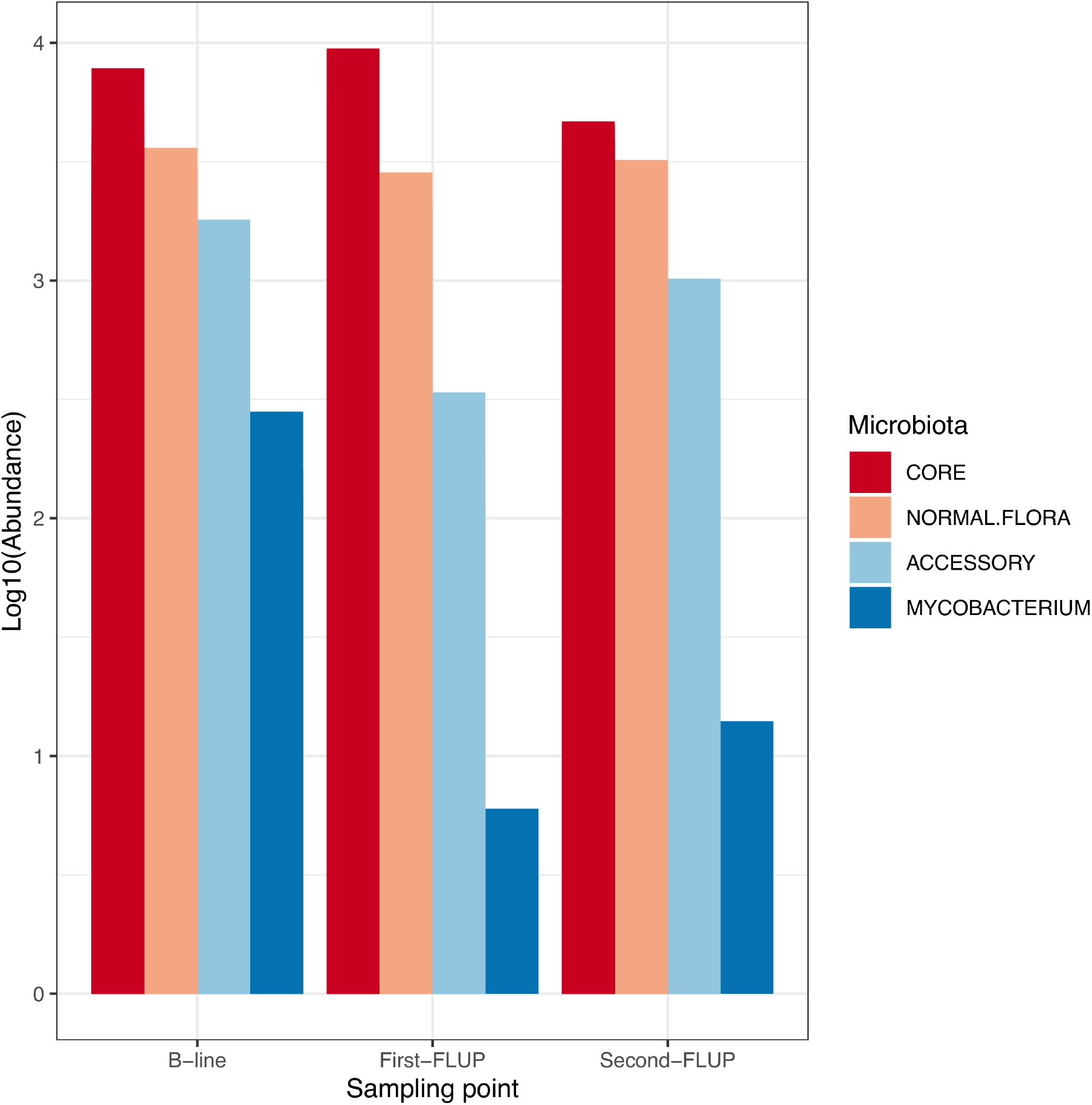
Changes in proportion of richness attributed to the core microbiota, accessory microbiota, oral-disease associated microbiota and *Mycobacterium*.

Furthermore, in-depth investigation of the abundance of the microbiota components (core, accessory and oral-disease associated) showed subtle differences (**Figure 4**). At baseline, there were two groups of patients i.e. clusters ii & iii (**Fig 4, panel A**) with a comparatively depleted core but these patients had a relatively high bacillary load i.e. 100-200+ colony forming units (CFUs). Although not exclusive, clusters i & iv (**Fig 4, panel A**) were predominated by patients who had commenced treatment. We also examined signatures of change in genera associated with oral-disease and accessory microbiota (**Fig 4, panels B** and **C**); we observed clustering with varying levels of depletion of genera associated with oral-pathology, with cluster iv mapping to sampling points associated with treatment follow-up (**Fig 4, panel B**). This observation was not seen with accessory microbiota, suggesting that this microbial component is less discriminatory. We observed that the accessory component was most dominant at baseline with enrichments of genera like *Bergeyella, Lactobacillus, Actinobacillus* and *Johnsonella* in certain patient groups during treatment. At month 2 posttreatment commencement, the accessory size and composition was significantly depleted. To further investigate this change in biomass and families attributed, we used Quasi-Poisson model to identify 11 differentially abundant families at baseline i.e. *Streptococcaceae, Neisseriaceae, Prevotellaceae* and *Peptostreptococcaceae*. These families also represent core membership described above and their differential abundance suggests dysbiosis at this point. Beyond month 2, the number of differentially abundant families reduced to seven. Furthermore, we noted that the abundance of core members significantly increased among patients with a high body mass index (BMI) compared to patients with low BMI by month 2 (**Figure 6**). We also noted that genera belonging to families *Tannerellacaeae* and *Leptotrichiacae* were exclusively differentially abundant after initiating therapy. Overall, this model shows that anti-TB therapy is associated with a reduction in microbial abundance/biomass with the largest change occurring at month 2 posttreatment commencement.

**Figure 4:**
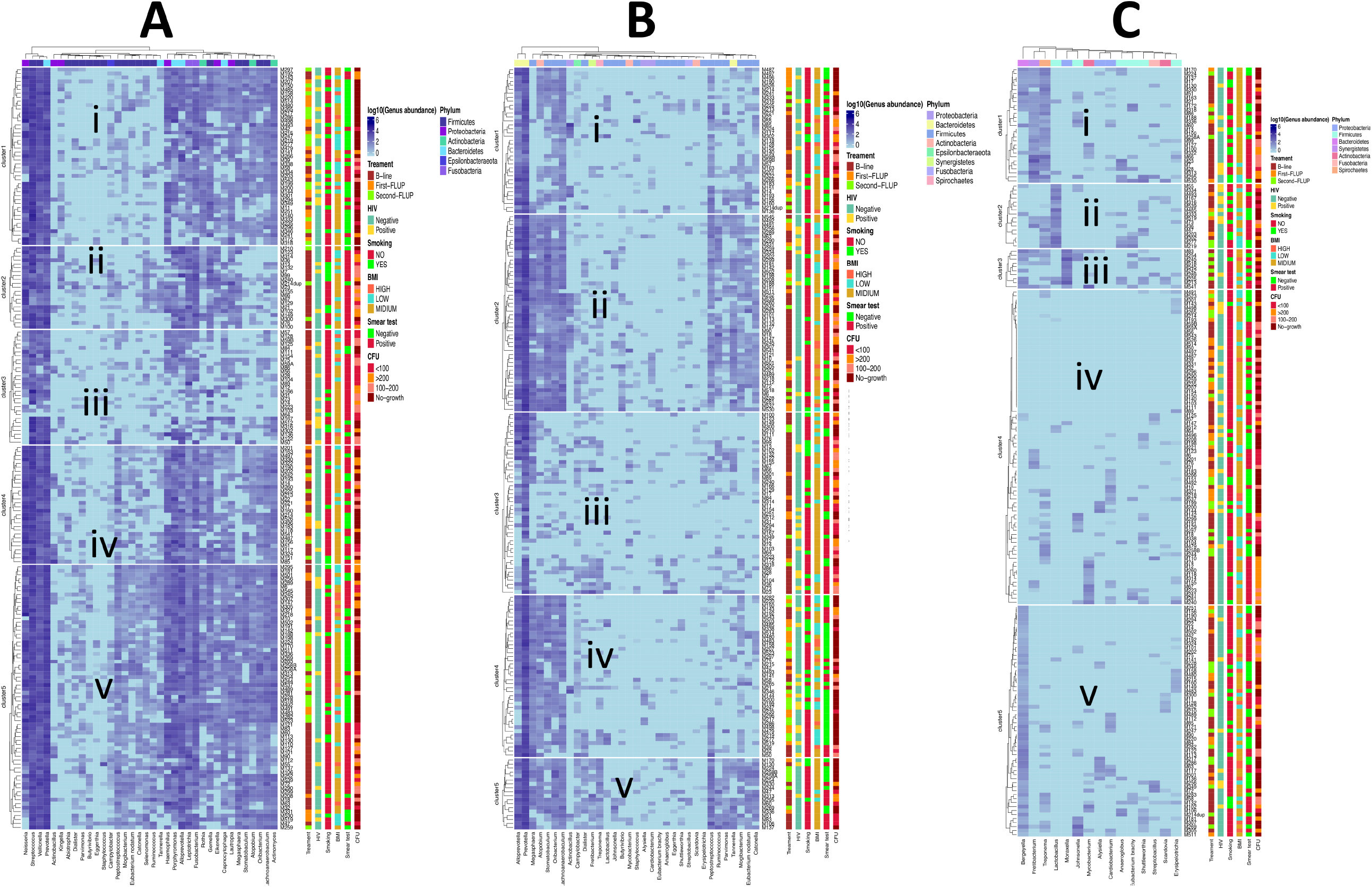
Changes in composition and abundance of the core microbiota (panel A), normal flora (panel B) and accessory microbiota during treatment (panel C). The clustering i-v shows patient groupings some of which correspond to clinical diagnostics like sputum smear microscopy results, bacillary load (CFU), HIV status and body mass index (BMI).

**Figure 6:**
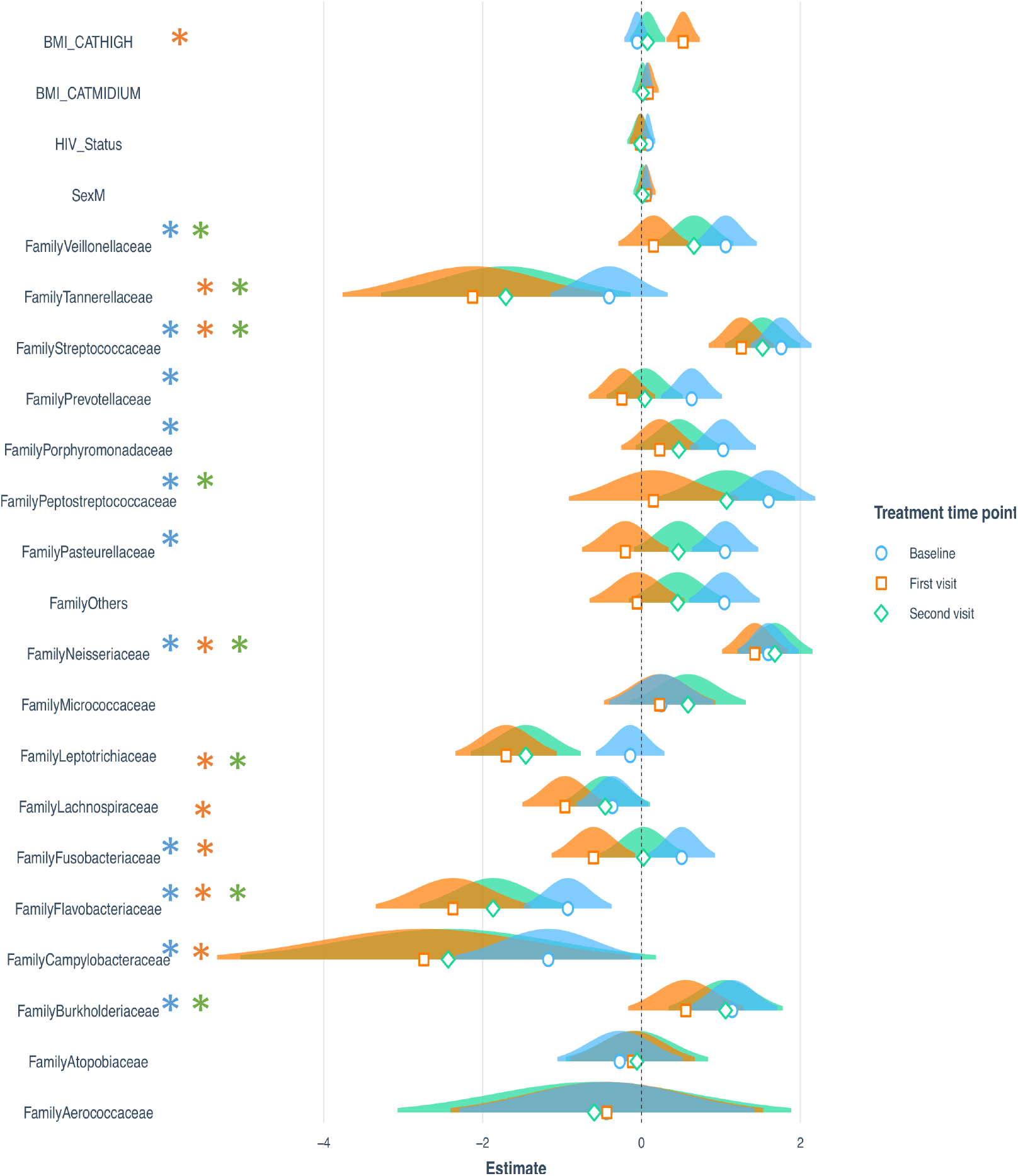
Changes in biomass using Poisson regression model. Blue, green and orange colors depict estimates of biomass at Baseline, month 2 (first visit) and month 5 (second visit) after initiating therapy. The pseudo R2 = 0.67, 0.95, and 0.94 for models, respectively. ***p <0.001; **p<0.01; *p<0.05.

### Community co-occurrence patterns and variance

Analysis of co-occurrence networks at baseline showed that the entire sputum microbiota community is linked by non-random relationships (p<0.05), **Fig**.**7 panel C** (‘non-random relationships’ refer to statistically significant patterns observed after applying a false discovery rate, FDR see Methods). Indeed, this was also seen at month 2 posttreatment commencement. Interestingly, at month 5 posttreatment commencement the proportion of random co-occurrence patterns was 70%. Co-occurrence network analysis of sputum samples at baseline revealed a community named × (**Fig**.**7 panel A**) predominated by genera belonging to accessory microbiota e.g. *Sulfitobacter, Pyramidobacter*, Acidiphilium, *Olivibacter* etc. By month 2, community × had shrunk in size but the co-occurrence network was still predominated by accessory microbiota. Indeed, there was a dramatic reduction in variance change (∼4%) at month 2 posttreatment commencement attributed to 35 genera, none of which were similar to those observed at baseline although five (14%) of these belonged to the core microbiota (**Table S2**). The largest change in variance at baseline (15%) was attributed to 10 genera –*Roseomonas, Proteobacterium, Acidocella, Spirochaeta, Olivibacter, Stenotrophomonas, Blautia, Desulfobulbus, Parasegetibacter* and *Terrimonas*, all of which belonged to accessory microbiota (**Figure 7, panel B** and **Table S2**). In contrast, at month 5 posttreatment commencement we observed up to 25% variance change attributed to 18 genera, 75% of which belonged to the core microbiota (**Figure 7 panel B** and **Table S2**).

**Figure 7:**
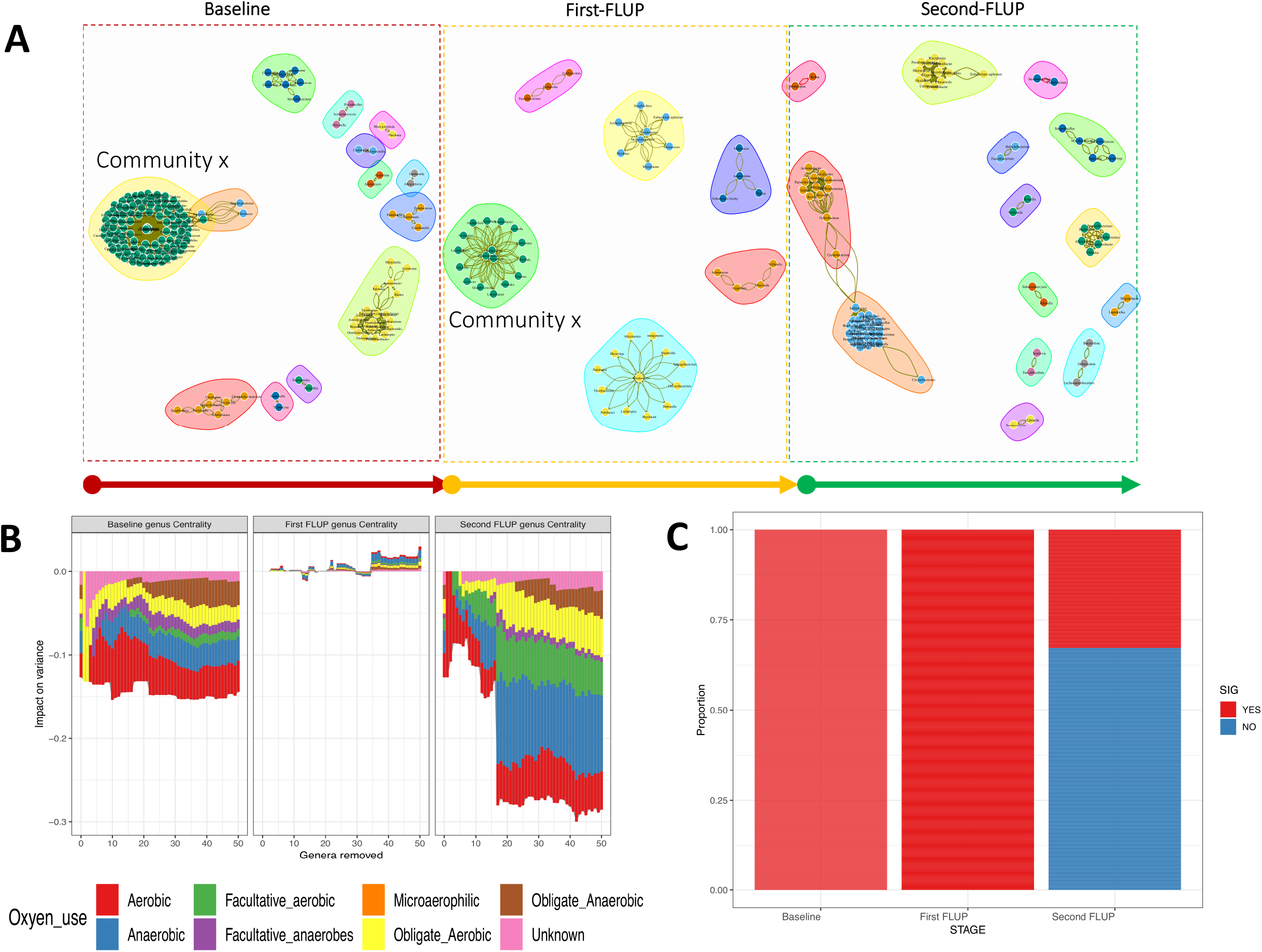
Shows changes in co-occurrence network patterns, variance and entropy. **Panel A** depicts remodeling of co-occurrence network during treatment. We highlight a community ‘x’ whose membership appears to change 2 months after initiating therapy. **Panel B** depicts change in variance attributed to genera with the highest centrality degree in co-occurrence network. Color coding represents oxygen utilization. **Panel C** depicts changes in proportion pairwise co-relations that are statistically significant. This was used as a measure of entropy i.e. 100% statistically significant (non-random) and random representing lack of entropy and high entropy, respectively. B-line, Baseline before initiating therapy; First-FLUP, First follow up (month 2); Second FLUP, Second follow up (month 5).

Normal flora significantly contributes to maintenance of the integrity of several anatomical sites in the body. Core microbiota as characterised in this study is a close approximation of the normal flora of the oral cavity and lung^11^. Understanding how this microbiota component is affected by anti-TB drugs is central to how it can be leveraged to improve diagnostics and treatment response monitoring^23^. We have shown in this study that on average a TB patient had a core microbiota size of 34 genera at any given time, which remains stable during treatment. Although the core size did not change, the relative abundance of its constituents changed for example, a significantly low abundance of *Mycobacterium* at month 2 (which is expected as the administered drugs target this genus). Indeed at baseline before initiating treatment, *Mycobacterium* was a member of the core and it accounted for 1 in 500 bacteria in sputum. By months 2 and 5, the proportion of *Mycobacterium* fell to 1 in 5,000 and 1 in 10,000 respectively, and its membership shifted to accessory microbiota. This observation, coupled with clustering of patients by Ziehl-Neelsen (ZN) sputum smear status, is in line with the importance of sputum as a sample of choice in TB diagnostics.

Furthermore, the observed change in abundance of the other members of the core microbiota i.e. genera like *Neisseria, Veillonella, Fusobacterium, Streptococcus, Actinobacteria, Capnocytophaga, Stomatobaculum* and *Prevotella* suggests dysbiosis^11,16^. Abnormality in microbiota composition and abundance (i.e. dysbiosis) is associated with dysregulation of the immune response, which alters the environment in favour of invading bacteria^10^ The observed high biomass of the accessory microbiota supports the notion that dysbiosis promotes proliferation of bad bacteria^10^. This accessory microbiota included genera like *Actinobacillus, Bergeyella* and *Fretibacterium*. On the other hand, genera constituting the core microbiota e.g. *Veillonella, Streptococcus, Peptostreptococcus, Porphyromonas, Neisseria, Pasteurella* and *Prevotella* were highly abundant before initiating therapy and such differential abundance of members of the core has been observed before^15,16^. Within 2 months after initiating therapy, we noted a general reduction in total microbiota abundance. This may not be surprising because rifampicin, one of the first-line anti-TB drugs administered to patients, has broad-spectrum of activity^11^ hence the overwhelming reduction in abundance of the microbiota. Even after controlling for factors like sex, HIV status and BMI, this marked change in abundance remained. This effect was more pronounced among genera that were abundant before initiating treatment, as well as *Tanerella, Leptotrichia, Lachnospira* and *Fusobacterium*, and it was also associated with a log reduction in abundance of the accessory microbiota.

In agreement with previous reports on effects of antibiotics on the microbiota^24^, we have shown that anti-TB treatment is associated with constriction in sputum microbiota community variance, lack of entropy and general reduction in biomass specifically that of accessory microbiota. The increase in community variance and entropy associated with 0.5 log increase in accessory microbiota at month 5 posttreatment commencement likely signals a gradual return to normal of the microbiota composition. We observed mainly positive rather than negative correlation, the latter being observed only at month 5 posttreatment commencement; this in addition to an increase in entropy could indeed signify sputum microbiota remodelling towards a normal/healthy state^25^. Overall, these findings suggest that accessory microbiota augments sputum microbiota dynamics but the pathobiological impact of this is currently unknown.

### Clinical relevance of findings for TB endemic settings with high incidence of HIV

As sputum is key in the diagnosis of pulmonary TB disease, investigation of sputum microbiota dynamics can inform efforts aiming to improve treatment response monitoring but this phenomenon is largely unexplored in Africa. In this study, sputum microbiota characteristics ably grouped patients as baseline (before initiating therapy) or treatment follow-up (months 2 or 5) and certain microbiota attributes also mapped to common diagnostics like AFB smear staining. Therefore, there is potential for this analysis to inform minimum detection levels for common TB diagnostics. We propose a framework (see **Figure 8**) for standardizing such methods for TB clinical metagenomics in the developing countries. The proposed framework combines a) sputum microbiota structure, b) taxonomic characteristics, and c) microbial network co-occurrence characteristics to provide holistic insight into microbial dynamics during therapy. This is critical because sputum only approximates the lung microbial distribution^17^, so relying on mere presence or absence of genera might be a misleading comparator.

**Figure 8:**
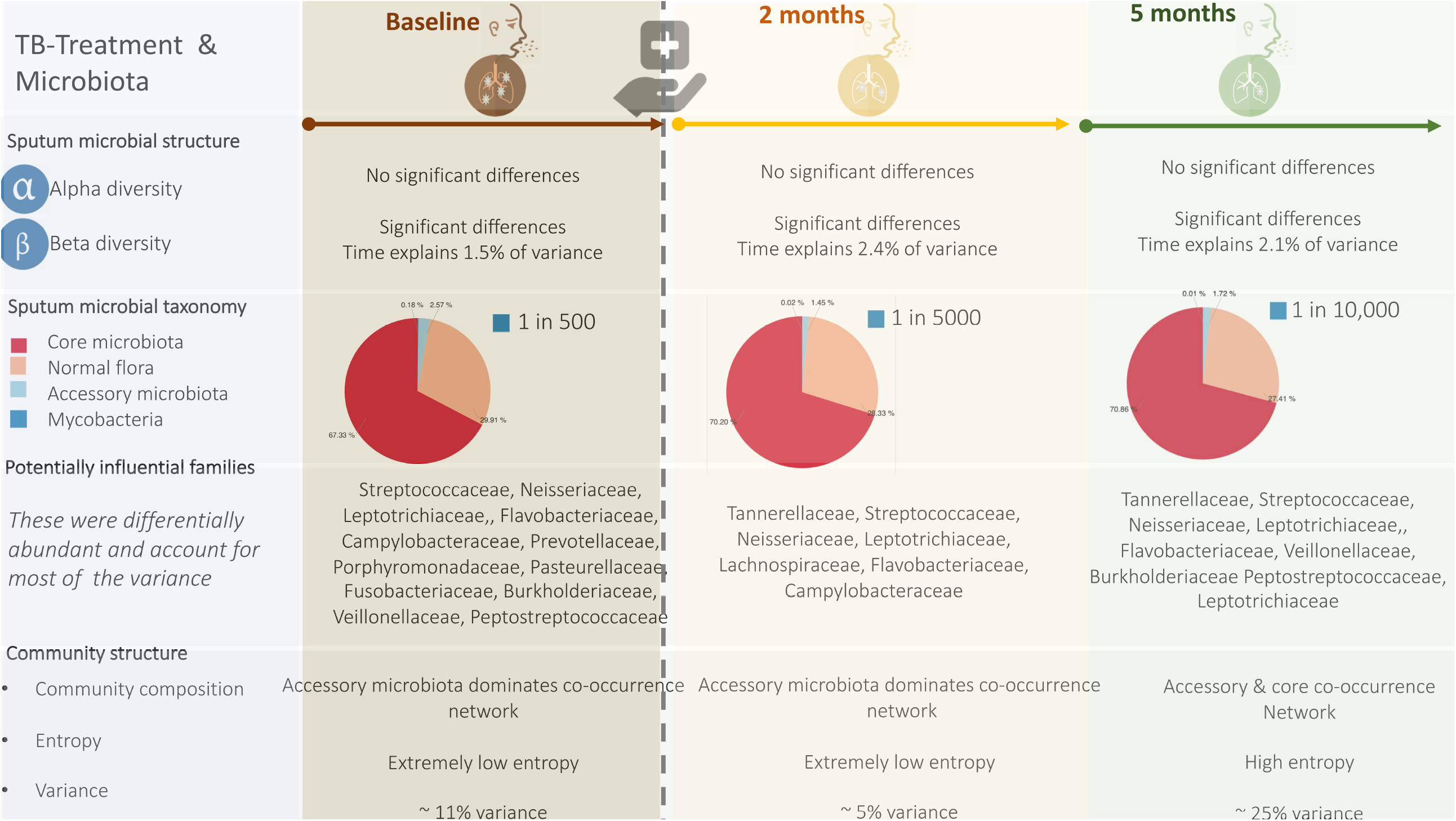
Proposed framework for characterizing sputum microbiota during anti-TB therapy in TB endemic settings with high rates of HIV infection. This model emphasizes use of microbiota structure, taxonomic characteristics, and co-occurrence networks including its variance and entropy.

## Conclusions

Sputum microbiota of treatment-naïve TB patients has microbial signals associated with first-line anti-TB therapy. Overall, TB treatment with first-line drugs does not change the size but the abundance of the core microbiota. In contrast, the size, composition and abundance of the accessory microbiota to which *Mycobacterium* belongs changes significantly during treatment. Also, treatment constricts microbial community variance and entropy, which recovers at month 5 posttreatment commencement. Taken together, this demonstrates the potential utility of sputum microbiome to inform treatment response monitoring strategies and refining diagnostics in developing countries like Uganda.

## Methods

### Study setting, patients and samples

This longitudinal study was conducted at Mulago National Referral Hospital in Kampala, Uganda, between 2016 and 2018. Mulago is the largest public hospital in Uganda with 1,500 beds, a TB clinic and a multidrug resistant (MDR)-TB treatment centre for the country. Around 5,000 TB patients are treated at the clinic every year, of whom one third are previously treated patients^26^. We randomly enrolled 120 treatment-naïve TB patients 18 years and older, collected sputum and profiled them before initiating anti-TB therapy which formed the baseline for the study. Pulmonary TB disease was clinically diagnosed by a Pulmonologist at the TB clinic and confirmed with the Xpert MTB/RIF test on sputum, sputum smear microscopy (by ZN staining for AFB) or LJ culturing at the BSL-3 Mycobacteriology Laboratory, Makerere University College of Health Sciences. We collected additional sputum samples from the patients during treatment follow-up at months 2 and 5; we compared these to baseline samples to unravel the microbiota changes and dynamics after initiating therapy. To account for potential cofounders, we included a wide range of clinical and lifestyle variables. Sputum samples were collected with consent as part of routine clinical care however, to ensure consistency in quality and quantity of the sputum collected, sputum induction was performed by an expert Pulmonologist at the TB clinic as described previously^27^. Specimen containers with sputum were tightly closed and placed in plastic biohazard bags and immediately transported to the BSL-3 Mycobacteriology laboratory in ice-cool boxes for processing. Briefly, N-acetyl-l-cysteine-sodium hydroxide (NALC-NaOH) and RLT Plus lysis buffer (Qiagen) were added to the samples, vortexed and centrifuged at 3,000 g for 2 minutes to obtain pellets. Pellets were re-suspended in sterile phosphate buffered saline and aliquoted into 2 portions. One portion was used for ZN staining and culturing in the BACTEC-MGIT system and on LJ medium. Chromosomal DNA was extracted from the remaining portion of the pellet by using the AllPrep DNA/RNA Mini Kit (Qiagen).

### DNA sequencing and sequence analysis

High throughput DNA sequencing targeting the V3-V4 region of the *16S rRNA* gene in sputum DNA was performed at the Integrated Microbiome Resource (IMR), Dalhousie University Canada (https://imr.bio)^28,29^ on MiSeq platform (illumina Inc.). Sequence reads were processed by using the Quantitative Insights into Microbial Ecology version 2 software (QIIME-2)^30^. Briefly, raw paired-end sequence reads and metadata were combined to generate a QIIME-2 object (artefact) for subsequent analysis^30^. Paired sequence reads were filtered for quality, dereplicated, chimeras removed and denoised by using DADA2^30^. This generated amplicon sequence variant (ASV) tables (previously known as operational taxonomic unit [OUT] tables) and representative sequences (in this study, the terms “ASVs” and “OTUs” are used interchangeably). The OTU tables were used to estimate alpha and beta diversity indices at an OTU minimum depth threshold of 2,000. This was followed by a step to identify potential contamination. Alpha diversity indices included observed OTUs and Shannon^31^. Beta diversity was estimated by using Bray-Curtis, weighted and unweighted uniFrac distances^30^. Phylogenetic analysis was inferred with phyloseq package v1.26.0^32^ and Metacoder v0.3.2-based microbiome analysis in R statistical package (v3.5.1)^14^ during beta diversity estimation. This was done by aligning the representative sequences, filtering out non-informative sites and generating a rooted tree using MAFFT and FASTTREE^30^ software. The taxonomic classification of OTUs was achieved by using a naïve Bayes classifier trained on the most recent SILVA database at 97% similarity^33^. First, the training dataset was extracted by using primers used for sequencing the samples^28^, and the resultant dataset used to train the classifier for taxonomically assigning the OTUs. Visualisation of taxonomic abundance was done by using ggplot2 in R. Phylogenetic abundance shifts of microbiota were interrogated by using Metacoder v0.3.2^32^. Additional post-hoc statistical analysis was done in R using phyloseq package v1.26.0^31^.

### Microbiota community structure and clinical covariates

To assess the explanatory power/effect size of variables for changes in community structure, we used sampling time at months 2 and 5 as proxy for monitoring anti-TB treatment. We performed a constrained analysis of distance-based redundancy; in this case we used Bray-Curtis as the outcome variable and sampling point and ZN sputum smear staining as the explanatory variables. To estimate the cumulative effect size of all our explanatory variables, we used PERMANOVA models with adonis function (9,999 permutations) in phyloseq v1.26.0^32^ to identify microbiota structure-associations. In addition, we compared Shannon diversity index and Richness using ANOVA and Kruskal Wallis depending on normality^31^.

### Characterising core and accessory microbiota

To determine the core and accessory microbiota, we used the microbiome package in R https://github.com/microbiome/microbiome. To track changes in the core as proportion of richness across sampling points, we used genera that were common to all cores for the three sampling points i.e. baseline, months 2 and 5.

### Co-occurrence networks analysis

To identify genera that co-occur at each sampling point, we used pairwise correlation method in the microbiome package v1.4.2 in R. First, we generated a genus-level spearman correlation matrix with mean abundances. Note that we applied a false discovery rate (FDR, Benjamini–Hochberg–Yekutieli) and filtered the adjusted p-value at 0.005 to limit spurious associations^34^. We then pruned the matrix using correlation coefficient (ρ) <=0.7 and >=-0.4. The resultant matrix was converted into a directed-network object from which communities were extracted and visualised in igraph package v1.2. To investigate changes in entropy i.e. if the co-occurrence patterns observed were non-random (non-random are the statistically significant patterns after applying FDR), we tracked the proportion of Random and Non-random patterns before and during therapy.

### Impact on total variance

To identify the influential genera (individual or groups of genera), we performed principal coordinate analysis (PCoA) and quantified total variance. Here total variance was the sum of variance explained by components 1-5 in a PCoA. Then, we sequentially removed the top 50 genera ranked by their network centrality (betweenness degree) in the co-occurrence network to compute the change in variance. The change in variance and attributable genera were plotted as a bar plot in ggplot2 coloured by oxygen utilisation (aerobic, anaerobic, etc.). The oxygen utilisation in this case was used as a functional community identifier based on the genera’s biological process Go-term in UniProt https://www.uniprot.org

### Poisson regression model for biomass

To assess changes in biomass and identify the associated clinical and lifestyle factors, we developed a Poisson regression model in glm using lme4 package in R. Here we assessed abundance at family level to minimise the number of covariate patterns. The families selected corresponded to core membership across the study period to ensure equal representation across the sampling points. Since the biomass variance was vastly different from the mean abundance, we used the quasi Poisson model. Here we developed a model with the same explanatory variables for each of the three sampling points and compared model estimates to determine the impact of TB therapy on sputum biomass.

### Ethical approval and consent to participate

Ethical approval was provided by the Research and Ethics Committee of the School of Biomedical Sciences, Makerere University College of Health Sciences (reference #s SBS381/SBS542). Written informed consent was obtained from all the participants prior to enrolment into the study. All study methods were performed in accordance with the relevant guidelines and regulations.

## Data Availability

All data generated or analysed during this study are included in this published article [and its supplementary information files].

## Acknowledgements

We thank: Professor Alison Elliott of Makerere University Infection & Immunity Programme (MUII-Plus) at the MRC/UVRI & LSHTM Uganda Research Unit, Entebbe, for useful comments on the manuscript; staff at the Mulago Hospital TB treatment centre and the BSL-3 Mycobacteriology laboratory at Makerere University for technical assistance; Harriet Nakayiza & Geraldine Nalwadda (Makerere University College of Health Sciences), and Joshua Mandre (MUII-plus, Entebbe) for administrative support.

This project is part of the EDCTP2 programme supported by the European Union (grant number TMA2018CDF-2357-MTI-Plus). The project was also supported in part by the DELTAS Africa Initiative (grant # 107743/Z/15/Z), the Africa Centre of Excellence in Materials, Product Development & Nanotechnology (MAPRONANO) (Project ID Number: P151847, IDA Number: 5797-UG), and the Erasmus Mobility Grant. The DELTAS Africa Initiative is an independent funding scheme of the African Academy of Sciences (AAS), Alliance for Accelerating Excellence in Science in Africa (AESA), and supported by the New Partnership for Africa’s Development Planning and Coordinating Agency (NEPAD Agency) with funding from the Wellcome Trust (Grant no. 107743) and the UK Government. The funders had no role in study design, data collection and analysis, decision to publish, or preparation of the manuscript.

## Author contributions statement

DPK & AM conceived the study, analysed and interpreted the data and wrote the manuscript. SN & AO recruited study participants and collected sputum samples from the patients. MM, FN, EK, WS & FKA performed the molecular and microbiological procedures. DPK, LN, WS & MLJ supervised the study protocol. MM performed part of the bioinformatics analysis (under supervision of DPK, AM & LN) in partial fulfilment of the requirements for the award of the degree of Master of Science in Immunology & Clinical Microbiology of Makerere University. All authors read and approved the final manuscript.

## Additional information

### Data Availability

All data generated or analyzed during this study are included in this published article (and its Supplementary Information files). The raw sequence data was deposited with links to BioProject accession number PRJNA564562 in the NCBI BioProject database https://www.ncbi.nlm.nih.gov/bioproject/

## References

1 Organization, W. H. & Organization, W. H. The top 10 causes of death. 2014. Fact sheet (2018).

2 Organization, W. H. Global tuberculosis report 2016. 2016. Google Scholar, 214 (2018).

3 Tuberculosis, U. N. & Programme, L. C. (2017).

4 Theron, G. et al. Feasibility, accuracy, and clinical effect of point-of-care Xpert MTB/RIF testing for tuberculosis in primary-care settings in Africa: a multicentre, randomised, controlled trial. The Lancet 383, 424–435 (2014).

5 Wipperman, M. F. et al. Antibiotic treatment for Tuberculosis induces a profound dysbiosis of the microbiome that persists long after therapy is completed. Scientific reports 7, 1–11 (2017).

6 WHO. (World Health Organization Geneva, 2017).

7 Rockwood, N., du Bruyn, E., Morris, T. & Wilkinson, R. J. Assessment of treatment response in tuberculosis. Expert Rev Respir Med 10, 643–654, doi:10.1586/17476348.2016.1166960 (2016).

8 Ambreen, A., Jamil, M., Rahman, M. A. u. & Mustafa, T. Viable Mycobacterium tuberculosis in sputum after pulmonary tuberculosis cure. BMC Infectious Diseases 19, 923, doi:10.1186/s12879-019-4561-7 (2019).

9 Su, W.-J., Feng, J.-Y., Chiu, Y.-C., Huang, S.-F. & Lee, Y.-C. Role of 2-month sputum smears in predicting culture conversion in pulmonary tuberculosis. European Respiratory Journal 37, 376–383, doi:10.1183/09031936.00007410 (2011).

10 Consortium, I. H. i. R. N. The integrative human microbiome project. Nature 569, 641–648 (2019).

11 Hong, B.-Y. et al. Microbiome changes during tuberculosis and antituberculous therapy. Clinical microbiology reviews 29, 915–926 (2016).

12 Namasivayam, S., Sher, A., Glickman, M. S. & Wipperman, M. F. The microbiome and tuberculosis: early evidence for cross talk. MBio 9, e01420–01418 (2018).

13 Pechal, J. L., Schmidt, C. J., Jordan, H. R. & Benbow, M. E. A large-scale survey of the postmortem human microbiome, and its potential to provide insight into the living health condition. Scientific reports 8, 1–15 (2018).

14 Handelsman, J., Rondon, M. R., Brady, S. F., Clardy, J. & Goodman, R. M. Molecular biological access to the chemistry of unknown soil microbes: a new frontier for natural products. Chemistry & biology 5, R245–R249 (1998).

15 Cheung, M. K. et al. Sputum microbiota in tuberculosis as revealed by 16S rRNA pyrosequencing. PLoS One 8 (2013).

16 Krishna, P., Jain, A. & Bisen, P. Microbiome diversity in the sputum of patients with pulmonary tuberculosis. European Journal of Clinical Microbiology & Infectious Diseases 35, 1205–1210 (2016).

17 Garcia, B. J. et al. Sputum is a surrogate for bronchoalveolar lavage for monitoring Mycobacterium tuberculosis transcriptional profiles in TB patients. Tuberculosis 100, 89–94 (2016).

18 Botero, L. E. et al. Respiratory tract clinical sample selection for microbiota analysis in patients with pulmonary tuberculosis. Microbiome 2, 29 (2014).

19 Luo, M. et al. Alternation of gut microbiota in patients with pulmonary tuberculosis. Frontiers in physiology 8, 822 (2017).

20 Wu, J. et al. Sputum microbiota associated with new, recurrent and treatment failure tuberculosis. PloS one 8 (2013).

21 Falony, G. et al. Population-level analysis of gut microbiome variation. Science 352, 560–564 (2016).

22 Astudillo-García, C. et al. Evaluating the core microbiota in complex communities: a systematic investigation. Environmental microbiology 19, 1450–1462 (2017).

23 Garcia, C. et al. High frequency of diarrheagenic Escherichia coli in human immunodeficiency virus (HIV) patients with and without diarrhea in Lima, Peru. Am J Trop Med Hyg 82, 1118–1120, doi:10.4269/ajtmh.2010.09-0460 (2010).

24 Langdon, A., Crook, N. & Dantas, G. The effects of antibiotics on the microbiome throughout development and alternative approaches for therapeutic modulation. Genome medicine 8, 39 (2016).

25 Mainali, K., Bewick, S., Vecchio-Pagan, B., Karig, D. & Fagan, W. F. Detecting interaction networks in the human microbiome with conditional Granger causality. PLoS computational biology 15, e1007037 (2019).

26 Bwanga, F., Haile, M., Joloba, M. L., Ochom, E. & Hoffner, S. Direct nitrate reductase assay versus microscopic observation drug susceptibility test for rapid detection of MDR-TB in Uganda. PLoS One 6 (2011).

27 Bell, D., Leckie, V. & McKendrick, M. The role of induced sputum in the diagnosis of pulmonary tuberculosis. Journal of Infection 47, 317–321 (2003).

28 Comeau, A. M., Douglas, G. M. & Langille, M. G. Microbiome helper: a custom and streamlined workflow for microbiome research. MSystems 2 (2017).

29 Walters, W. et al. Improved bacterial 16S rRNA gene (V4 and V4-5) and fungal internal transcribed spacer marker gene primers for microbial community surveys. Msystems 1, e00009–00015 (2016).

30 Bolyen, E. et al. QIIME 2: Reproducible, interactive, scalable, and extensible microbiome data science. Report No. 2167-9843, (PeerJ Preprints, 2018).

31 McMurdie, P. J. & Holmes, S. phyloseq: an R package for reproducible interactive analysis and graphics of microbiome census data. PloS one 8 (2013).

32 Foster, Z. S., Sharpton, T. J. & Grünwald, N. J. Metacoder: an R package for visualization and manipulation of community taxonomic diversity data. PLoS computational biology 13, e1005404 (2017).

33 Quast, C. et al. The SILVA ribosomal RNA gene database project: improved data processing and web-based tools. Nucleic acids research 41, D590–D596 (2012).

34 Proctor, L. M. et al. The Integrative Human Microbiome Project. Nature 569, 641–648, doi:10.1038/s41586-019-1238-8 (2019).

